# Childhood vaccination coverage and equity impact in Ethiopia by socioeconomic, geographic, maternal and child characteristics

**DOI:** 10.1101/2020.01.03.20016436

**Authors:** Anne Geweniger, Kaja M. Abbas

**Author notes:** Corresponding author at: Center for Paediatrics and Adolescent Medicine, University Medical Center, Heiliggeiststr.1, 79106 Freiburg, Germany.

## Abstract

**Background:** Ethiopia is a priority country of Gavi, the Vaccine Alliance to improve vaccination coverage and equitable uptake. The Ethiopian National Expanded Programme on Immunisation (EPI) and the Global Vaccine Action Plan set coverage goals of 90% at national level and 80% at district level by 2020. This study aims to analyse basic vaccination coverage among children in Ethiopia and to estimate the equity impact by socioeconomic, geographic, maternal and child characteristics based on data from the Ethiopia Demographic and Health Survey 2016.

**Methods:** Basic vaccination coverage (1-dose BCG, 3-doses DPT-HepB-Hib, 3-doses polio, 1-dose measles vaccine) of 2,004 children aged 12-23 months was analysed. Mean coverage was disaggregated by socioeconomic (household wealth, religion, ethnicity), geographic (area of residence, region), maternal (maternal age at birth, maternal education, maternal marital status, sex of household head) and child (sex of child, birth order) characteristics. Concentration indices assessed wealth and education-related inequalities. Multiple logistic regression estimated associations between basic vaccination coverage and socioeconomic, maternal and child characteristics.

**Results:** National coverage for basic vaccinations was 39.7% in 2016. Single vaccination coverage ranged between between 53.2% (DTP3) and 69.2% (BCG). Wealth and maternal education related inequities were present for all vaccines. Children from richer households, urban regions, primary maternal education and male headed households were associated with higher vaccination coverage. The Ethiopia Mini Demographic and Health Survey 2019 reports national coverage for basic vaccinations at 43.3% with single vaccination coverage ranging between 57.8% (measles) and 74.2% (BCG).

**Conclusions:** Vaccination coverage has improved from 2016 to 2019, but remains below the coverage goals of the EPI. Low vaccination coverage is associated with poorer households, rural regions of Afar and Somali, no maternal education and female headed households. Targeted approaches are necessary to improve vaccination coverage among these population subgroups and equitable uptake of vaccines in Ethiopia.

## Introduction

Ethiopia had a total population of 109 million with gross national income per capita of US$ 790 in 2018 [1]. 14.9% of the total population are under the age of 5 years, with an infant mortality rate of 41 per 1000 live births and under-five mortality rate of 59 per 1000 births [2]. Vaccine preventable diseases are an important cause of childhood mortality with an under-five mortality of 25,970 deaths due to lower respiratory infections and 14,662 deaths due to diarrheal diseases in 2015 [3].

Vaccines are considered one of the most safe and cost-effective interventions to reduce childhood morbidity and mortality [4]. Globally, 2-3 million deaths are currently prevented by vaccination every year [5]. However, at present 19.4 million children under the age of one year do not receive basic vaccines, 60% of whom live in Angola, Brazil, Congo, Ethiopia, India, Indonesia, Nigeria, Pakistan, Philippines and Vietnam.

### Equity in vaccination coverage

Vaccination coverage is monitored as part of Sustainable Development Goal 3, “(to) ensure healthy lives and promote well-being for all at all ages”, which aims at ending preventable deaths of newborns and children under the age of 5 years by 2030, while the goal of SDG 10 is to “Reduce inequality within and among countries” [6]. Further, 14 of the 17 SDGs are related to the success of immunisation programmes in improving vaccination coverage and equitable uptake [7].

The Global Vaccine Action Plan 2011-2020 includes equitable access to immunisations among its six guiding principles and strategic objectives, advocating for the monitoring of equity indicators, such as the gap in vaccination coverage between the highest and lowest wealth quintiles [4]. Gavi, the Vaccine Alliance has facilitated in improving immunisation access in the poorest countries through public-private partnerships, and incorporates measures of vaccine equity relating to geographic distribution, wealth and maternal education in its 2016-2020 vaccine goal indicators [8].

The Ethiopian National Expanded Programme on Immunisation (EPI) aligns with the Sustainable Development Goals 2016-2030, the Global Vaccine Action Plan 2011-2020, and Gavi, the Vaccine Alliance in setting equity goals for health outcomes and vaccination coverage. The EPI and the Global Vaccine Action Plan have set coverage goals of 90% at national level and 80% at district levels by 2020.

### Expanded Programme on Immunisation in Ethiopia

Ethiopia’s EPI was launched in 1980, and the routine immunisation schedule comprises of six vaccine preventable diseases, namely measles, diphtheria, pertussis, tetanus, polio and tuberculosis. Ethiopia has received support from Gavi since 2002 [9], and is one of ten tier-1 Gavi priority countries with the lowest national coverage of three doses of the combined diphtheria, tetanus toxoid and pertussis vaccine (DTP3), a commonly used indicator for immunisation programme performance [10,11]. Rotavirus, pneumococcal and second dose of measles vaccines have been introduced in 2011, 2012 and 2019 respectively [12]. The comprehensive multi-year plan 2016-2020 of Ethiopia’s National Expanded Programme on Immunisation states its first objective is to “reach 90% national coverage and 80% in every district with all vaccines by 2020” [13].

### Study objective

The aim is to analyse basic vaccination coverage among children in Ethiopia and estimate the equity impact by socioeconomic, geographic, maternal and child characteristics using data from the Ethiopia Demographic and Health Survey 2016. Aggregate country-level analysis conceals the hidden inequities in vaccination coverage, and the disaggregated equity impact analysis of this study will be valuable to identify the underserved subpopulations and inform policies and practices to address the vaccination barriers specific to these subpopulations [14].

## Methods

### Survey data

The data source for this study is the Ethiopia Demographic and Health Survey 2016, of which the full survey dataset is available [15]. A survey report on the Ethiopia Mini Demographic and Health Survey 2019 was released in July 2019 containing key indicators but primary data is not yet available [12].

The DHS surveys provide estimates of key demographic and health indicators, covering population, maternal and child health issues with a special focus on marriage, sexual and reproductive health, child health, nutrition, and HIV/AIDS. The surveys collect vaccination coverage data of children aged up to 36 months who had received specific vaccines at any time before the surveys according to their vaccination card, mother’s report and health facility records. For vaccine coverage estimation, data of of 1929 children aged 12-23 months from the 2016 DHS survey were included in the analysis, resulting in a total number of 2004 children after taking into account stratification, cluster sampling and differences in selection probability of participants through sample weights.

### Vaccination coverage

We primarily assessed basic vaccination coverage as defined by the WHO guidelines -- one dose of Bacille Calmette-Guérin (BCG) vaccine, three doses of pentavalent vaccine against DTP-Haemophilus influenza type b-Hepatitis B, three doses of polio vaccine and one dose of measles vaccine at the age of 12-23 months [16]. The coverage of single vaccinations and age-appropriate vaccinations at 12-23 months according to the Ethiopian vaccination schedule was also assessed [17]. Age-appropriate vaccinations include all basic vaccinations (as per WHO guidelines) as well as three doses of pneumococcal conjugate vaccine (PCV 3) and two doses of rotavirus vaccine [16,18]. Receipt of three doses of the combined diphtheria, tetanus toxoid and pertussis vaccine (DTP3) is widely used as an indicator of immunisation programme performance and ability of families to repeatedly access immunisation services [11,19].

### Equity criteria

Based on the equity criteria according to the guidance on priority setting in health care (WHO GPS-Health), particularly those related to social groups, and the assessment of inequalities in childhood immunisation in ten Gavi priority countries by WHO, the following explanatory variables were chosen as relevant stratifiers for the measurement of inequalities in vaccination coverage: wealth index; maternal age at birth, maternal education and marital status; sex of household head; area and region of residence; ethnicity; religion; sex and birth order of the child [9,20,21].

### Equity impact analysis

The distribution of vaccination coverage by socioeconomic, geographic, maternal and child characteristics was analysed to compute absolute and relative equity metrics, and estimated the contribution of these characteristics to inequalities in vaccination coverage through logistic regression. Simple logistic regression was conducted to obtain unadjusted odds ratios for the association between each exposure variable and basic vaccination coverage. The adjusted associations between basic immunisation coverage as binary outcome variable and selected exposure variables were assessed through multiple logistic regression (see Appendix A1). The analysis was conducted using Stata statistical software [22] by taking into account stratification, cluster sampling and differences in selection probability of participants through sample weights (see Appendix A2).

### Ethics approval

This study was approved by the ethics committee (Ref 16846) of the London School of Hygiene & Tropical Medicine. All authors had full access to all the data in the study and final responsibility for the decision to submit for publication.

## Results

### Vaccination coverage

2004 children aged 12-23 months and their mothers are represented in the 2016 Ethiopia DHS after applying sample weights. The mean DTP3 vaccination coverage among these children at national level was 53.2%. Vaccination coverage for other single vaccines ranged from 49.1% for PCV to 69.2% for BCG vaccine. The vaccination coverage for basic vaccinations (1-dose BCG, 3-doses DPT-HepB-Hib, 3-doses polio, 1-dose measles vaccines) and age-appropriate vaccinations (basic vaccinations plus 3-doses PCV and 2-doses rotavirus vaccines) was 39.7% and 33.8% respectively (see Table 1).

**Table 1:** Vaccination coverage in Ethiopia at the national level and by rural and urban areas. Vaccination coverage among children aged 12-23 months in Ethiopia and disaggregated by rural and urban areas of residence (mean coverage and 95% confidence intervals). Basic vaccinations includes 1-dose BCG, 3-doses DPT-HepB-Hib, 3-doses polio, 1-dose measles vaccines. Age-appropriate vaccinations include vaccines included in basic vaccinations plus 3-doses PCV and 2-doses rotavirus vaccines.

| Vaccine | Coverage (%) |  |  |  |
| --- | --- | --- | --- | --- |
|  | National<br>(N = 2004) | Rural<br>(n<sub>1</sub> = 1772) | Urban<br>(n<sub>2</sub> = 232) | Urban-rural<br>difference<sup>1</sup> |
| BCG | 69.2<br>(65.5, 72.8) | 66.6<br>(62.6, 70.5) | 88.8<br>(80.9, 96.7) | 22.3<br>(13.5, 31.1) |
| DPT-HepB-Hib | 53.2<br>(48.9, 57.4) | 49.7<br>(45.2, 54.2) | 79.5<br>(68.2, 90.7) | 29.8<br>(18.7, 41.8) |
| Polio | 61.8<br>(57.2, 66.3) | 58.9<br>(54.0, 63.9) | 83.2<br>(72.0, 94.4) | 24.2<br>(12.1, 36.4) |
| Measles | 54.3<br>(50.2, 58.5) | 51.5<br>(47.1, 55.9) | 76.0<br>(63.6, 88.4) | 24.5<br>(11.4, 37.5) |
| Basic vaccinations | 39.7<br>(35.5, 43.9) | 36.4<br>(32.1, 40.7) | 65.0<br>(52.2, 77.9) | 28.6<br>(15.1, 42.1) |
| PCV | 49.1<br>(45.1, 53.1) | 46.0<br>(41.8, 50.2) | 72.9<br>(61.2, 84.6) | 26.9<br>(14.5, 39.3) |
| Rotavirus | 56.0<br>(52.0, 59.9) | 52.9<br>(48.8, 57.1) | 79.1<br>(66.4, 91.8) | 26.1<br>(12.8, 39.4) |
| Age-appropriate<br>vaccinations | 33.8<br>(30.0, 37.7) | 30.3<br>(26.4, 34.1) | 61.1<br>(48.3, 73.9) | 30.9<br>(17.6, 44.2) |

<sup>1</sup> $p < 0.001$ for the respective differences in mean vaccination coverage between rural and urban areas of residence.

The urban-rural differences in vaccination coverage were significant at the 1%-level with a pro-urban advantage. The absolute differences in coverage were higher for DTP3, basic and age-appropriate vaccinations at 29.5%, 28.6% and 30.9% respectively and lowest for BCG vaccination at 22.3%.

### Equity impact: Vaccination coverage disaggregated by socioeconomic, geographic, maternal and child characteristics

The inequities in basic vaccination coverage in Ethiopia disaggregated by socioeconomic, geographic, maternal and child characteristics are illustrated in Table 2 and Figure 1. With respect to household wealth status, basic vaccination coverage was lowest among the poorest quintile (23.2%) and highest in the richest quintile (64.2%). Children in the richest quintile had 6 times the odds of basic vaccination coverage compared with children in the poorest quintile (crude OR 5.93; 95% CI: 3.20-10.99). With respect to religion, basic vaccination coverage was relatively higher among Christian children at 49.4% in comparison to Muslim children at 27.4%, and with respect to ethnicity, it was relatively higher among Tigray children at 69.1% and lowest among Somali children at 24.8%. With respect to geographic characteristics, basic vaccination coverage among children in urban and rural areas were 65% and 36.4% respectively while it was highest in the capital Addis Ababa at 89.2% and lowest in the Afar region at 15.2%.

**Table 2:** Inequities in basic vaccination coverage in Ethiopia by socioeconomic, geographic, maternal and child characteristics. Inequities in basic vaccination coverage (1-dose BCG, 3-doses DPT-HepB-Hib, 3-doses polio, 1-dose measles vaccine) among children aged 12-23 months by socioeconomic (household wealth, religion, ethnicity), geographic (area of residence, region), maternal (maternal age at birth, maternal education, maternal marital status, sex of household head) and child (sex of child, birth order) characteristics, based on simple logistic regression estimates of crude odds ratios (mean and 95% confidence intervals).

| Characteristics | Population<br>in each<br>subgroup n<br>(N=2,004) | Mean<br>basic<br>coverage<br>(%) | Odds Ratio | p-value |
| --- | --- | --- | --- | --- |
| Household wealth (quintiles) |  |  |  |  |
| poorest (referent) | 504 | 23.2 |  | <0.0001 |
| poorer | 396 | 39.0 | 2.12 (1.29, 3.46) |  |
| middle | 450 | 37.3 | 1.97 (1.18, 3.28) |  |
| richer | 366 | 46.8 | 2.91 (1.76, 4.80) |  |
| richest | 288 | 64.2 | 5.93 (3.20, 10.99) |  |
| Religion |  |  |  |  |
| Muslim (referent) | 786 | 27.4 |  | <0.0001 |
| Christian | 1171 | 49.4 | 2.59 (1.78, 3.77) |  |
| Traditional and other | 47 | 3.9 | 0.11 (0.01, 0.83) |  |
| Ethnicity |  |  |  |  |
| Somali (referent) | 72 | 24.8 |  | <0.0001 |
| Amhara | 419 | 51.1 | 3.17 (1.71, 5.90) |  |
| Oromo | 824 | 26.6 | 1.10 (0.60, 2.03) |  |
| Tigray | 155 | 69.1 | 6.80 (3.52, 13.12) |  |
| other | 534 | 44.5 | 2.44 (1.34, 4.44) |  |
| Area of residence |  |  |  |  |
| rural (referent) | 1772 | 36.4 |  | 0.0001 |
| urban | 232 | 65.0 | 3.25 (1.80, 5.87) |  |
| Region |  |  |  |  |
| Afar (referent) | 20 | 15.2 |  | <0.0001 |
| Tigray | 152 | 68.7 | 12.28 (5.29, 28.47) |  |
| Amhara | 364 | 46.7 | 4.91 (2.16, 11.16) |  |
| Oromia | 881 | 26.2 | 1.99 (0.89, 4.43) |  |
| Somali | 76 | 23.6 | 1.73 (0.71, 4.20) |  |
| Benishangul | 21 | 61.7 | 9.03 (3.88, 21.02) |  |
| Southern Nations,<br>Nationalities & People | 419 | 47.4 | 5.04 (2.26, 11.21) |  |
| Gambela | 5 | 42.7 | 4.18 (1.76, 9.92) |  |
| Harari | 5 | 43.0 | 4.22 (1.80, 9.92) |  |
| Addis Ababa | 52 | 89.2 | 46.41 (18.18, 118.5) |  |
| Dire Dawa | 9 | 79.0 | 21.07 (8.32, 53.36) |  |
| Maternal age at birth (years) |  |  |  |  |
| 15-19 (referent) | 238 | 33.5 |  | 0.03 |
| 20-34 | 1425 | 42.4 | 1.46 (0.98, 2.18) |  |
| 35-49 | 341 | 33.1 | 0.98 (0.61, 1.58) |  |
| Maternal education |  |  |  |  |
| none (referent) | 1257 | 32.3 |  | <0.0001 |
| primary | 577 | 46.7 | 1.83 (1.36, 2.46) |  |
| secondary or higher | 170 | 70.9 | 5.11 (2.61, 9.99) |  |
| Maternal marital status |  |  |  |  |
| Not married or formerly married (referent) | 107 | 30.6 |  | 0.04 |
| Married, not residing with partner | 155 | 29.4 | 0.94 (0.42, 2.10) |  |
| Married and residing with partner | 1742 | 41.2 | 1.59 (0.86, 2.93) |  |
| Sex of household head |  |  |  |  |
| male (referent) | 1705 | 41.2 |  | 0.02 |
| female | 299 | 31.5 | 0.66 (0.59, 0.84) |  |
| Sex of child |  |  |  |  |
| male (referent) | 926 | 37.8 |  | 0.30 |
| female | 1078 | 41.4 | 1.16 (0.87, 1.55) |  |
| Birth order |  |  |  |  |
| 6<sup>th</sup> born or higher (referent) | 569 | 31.0 |  | 0.001 |
| First born | 372 | 47.7 | 2.03 (1.37, 3.00) |  |
| 2<sup>nd</sup>-3<sup>rd</sup> born | 604 | 44.2 | 1.76 (1.23, 2.53) |  |
| 4<sup>th</sup>-5<sup>th</sup> born | 459 | 38.2 | 1.38 (0.90, 2.10) |  |

**Figure 1:**
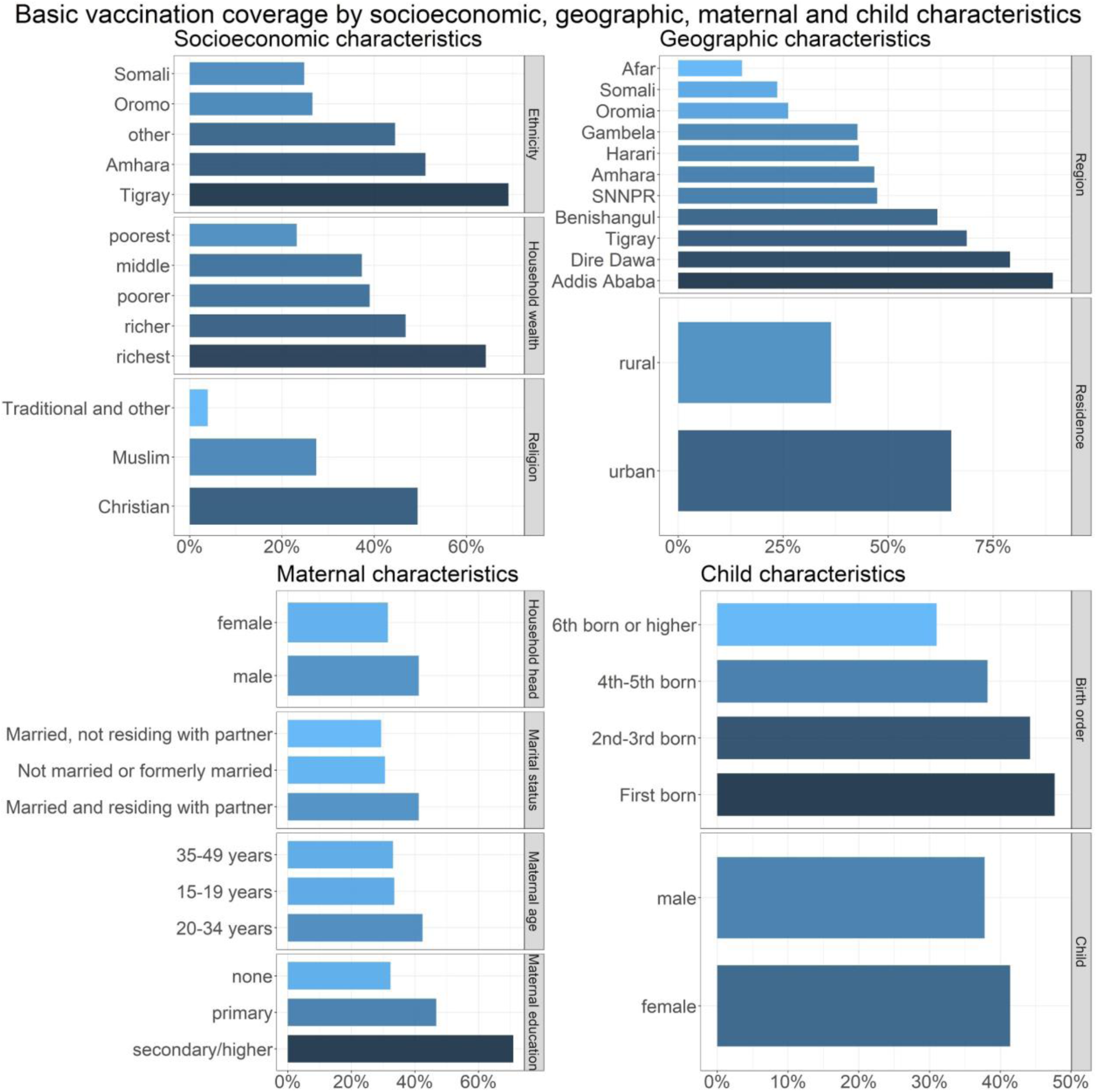
Vaccination coverage in Ethiopia among children aged 12-23 months by socioeconomic, geographic, maternal and child characteristics. Basic vaccination (1-dose BCG, 3-doses DPT-HepB- Hib, 3-doses polio, 1-dose measles vaccines) coverage in Ethiopia among children aged 12-23 months by socioeconomic (household wealth, religion, ethnicity), geographic (area of residence, region), maternal (maternal age at birth, maternal education, maternal marital status, sex of household head), and child (sex of child, birth order) characteristics.

With respect to maternal characteristics, basic vaccination coverage was lowest among children whose mothers were between 15-19 or 35-49 years old at the time of birth (33.5% and 33.1% respectively), had no education (32.3%), were not married or not residing with a partner (30.6% and 29.4% respectively), or who were living in a household with a female household head (31.5%). Children whose mothers had secondary or higher education had 5 times the odds of basic vaccination coverage compared to children whose mothers had no education (crude OR 5.11; 95% CI: 2.61-9.99). With respect to child characteristics, basic vaccination coverage was relatively highest among the first born children at 47.7%, and there were no sex-discrepancies in basic vaccination coverage.

### Wealth and maternal education related inequities in vaccination coverage

The vaccination coverage in Ethiopia have a pro-rich distribution with respect to both household wealth and maternal education, with relatively higher vaccination coverage among children from richer households and higher maternal education (Table 3 and Figure 2). The wealth-related concentration index varied from 0.22 for measles vaccination to 0.30 for pentavalent and age-appropriate vaccinations and the maternal education-related concentration index varied from 0.16 for measles vaccination to 0.23 for basic vaccinations. Thereby with respect to both household wealth and maternal education, we infer relatively higher equitable uptake of measles vaccines in comparison to other childhood vaccines in Ethiopia.

**Table 3:**
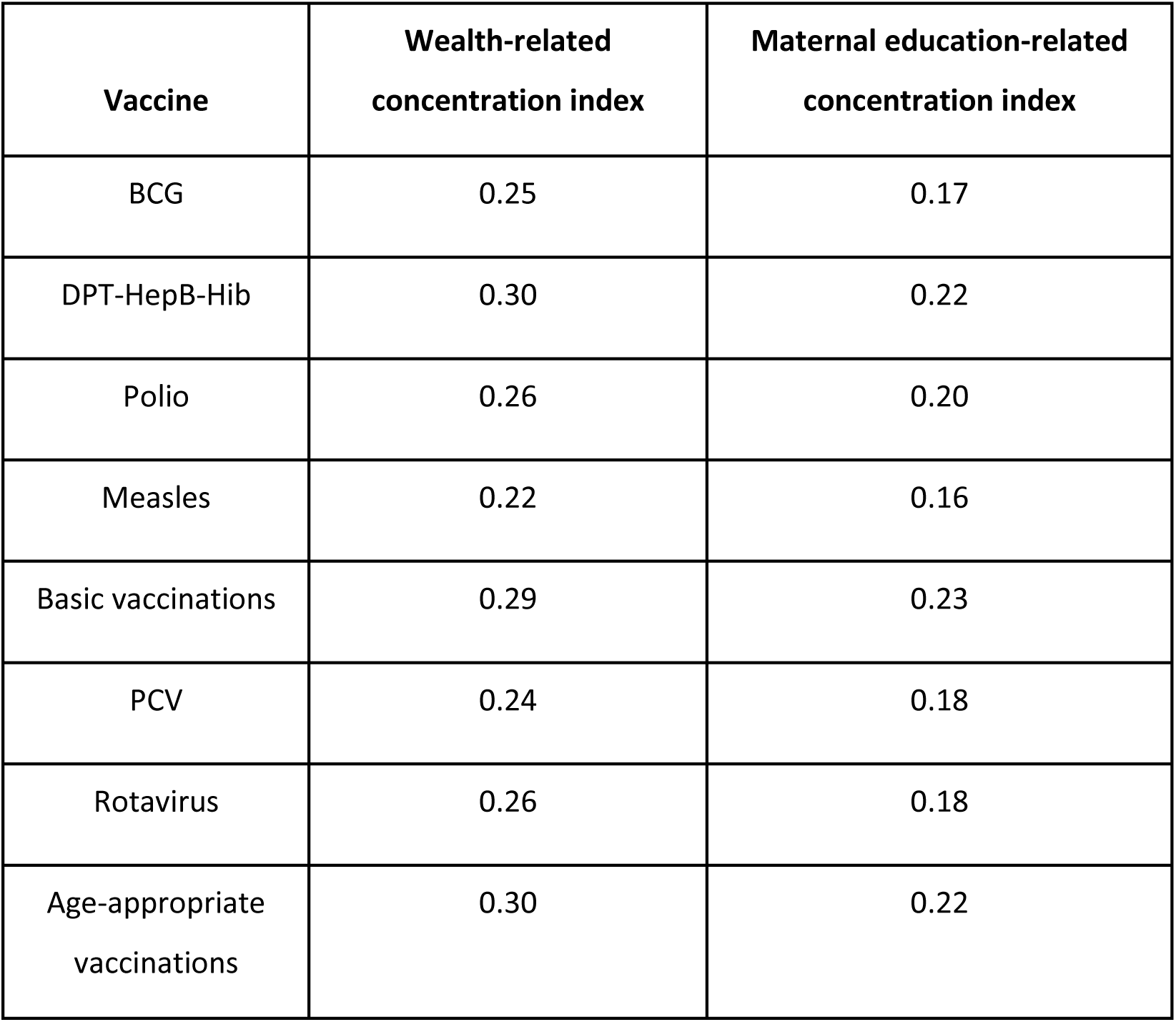
Wealth and maternal education related inequities in vaccination coverage for Ethiopia. Wealth and maternal education-related concentration indices for vaccination coverage in children aged 12-23 months, based on DHS 2016 survey. Basic vaccinations includes 1-dose BCG, 3-doses DPT-HepB-Hib, 3-doses polio, 1-dose measles vaccines. Age-appropriate vaccinations include vaccines included in basic vaccination plus 3-doses PCV and 2-doses rotavirus vaccines.

**Figure 2:**
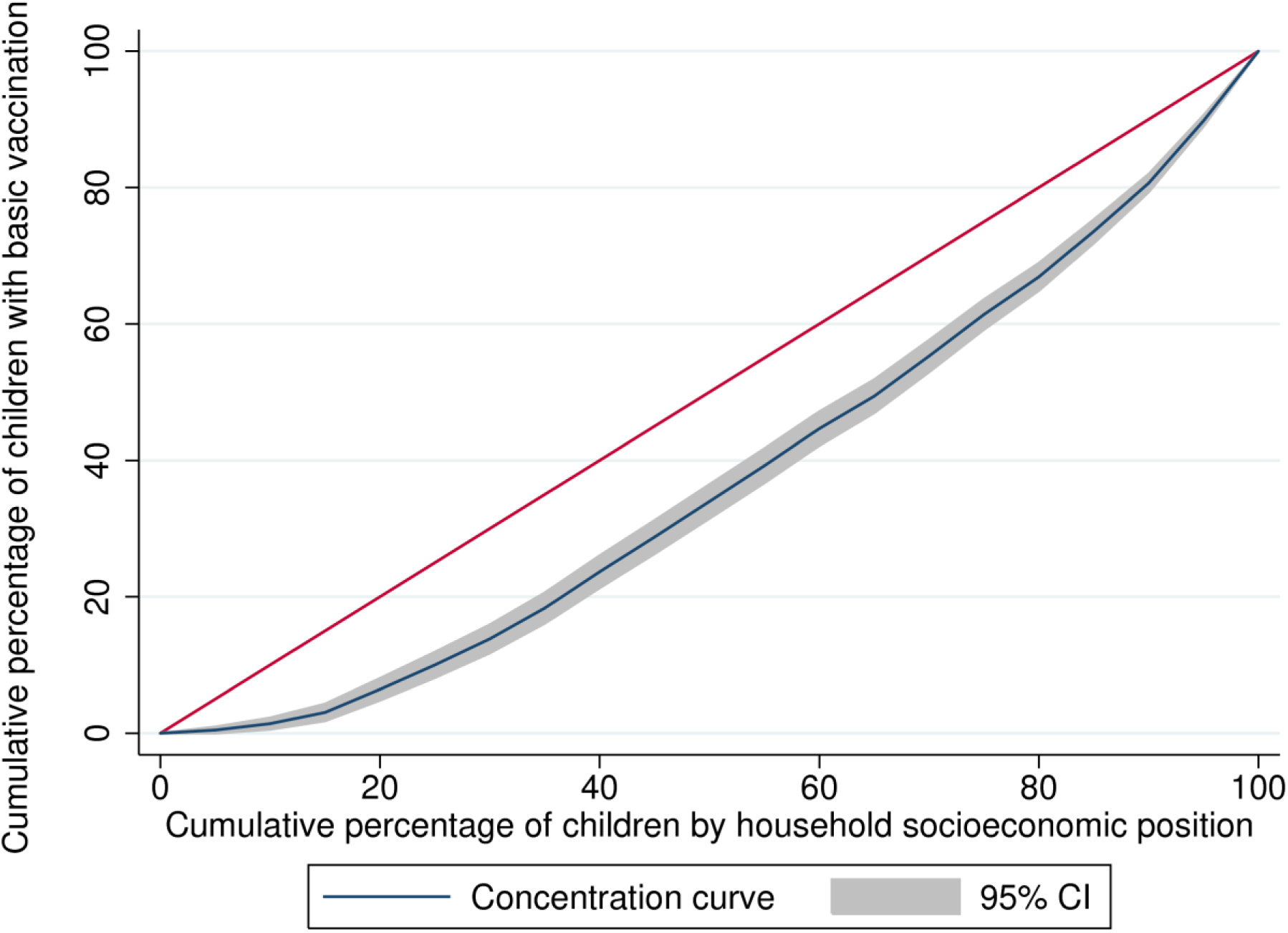
Wealth related inequity in vaccination coverage in Ethiopia. Concentration curve for basic vaccination (1-dose BCG, 3-doses DPT-HepB-Hib, 3-doses polio, 1-dose measles vaccines) coverage in children aged 12-23 months in Ethiopia by household wealth.

**Figure 3:**
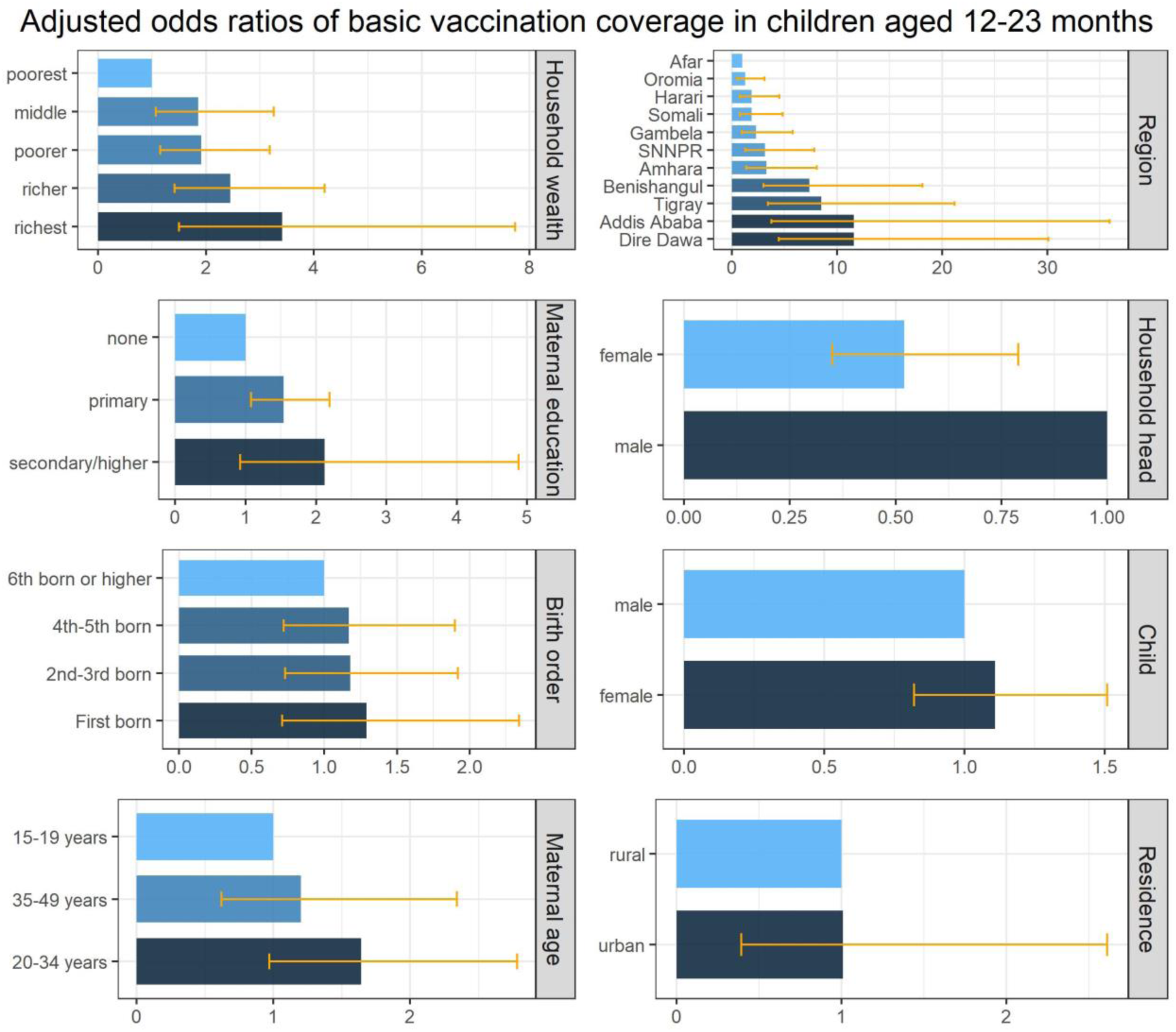
Inequities in vaccination coverage in Ethiopia associated with socioeconomic, geographic, maternal and child characteristics. Inequities in basic vaccination (1-dose BCG, 3-doses DPT-HepB-Hib, 3-doses polio, 1-dose measles vaccine) coverage among children aged 12-23 months associated with socioeconomic (household wealth), geographic (area of residence, region), maternal (maternal age at birth, maternal education, sex of household head) and child (sex of child, birth order) characteristics, based on multiple logistic regression estimates of adjusted odds ratios.

### Equity impact: Vaccination coverage associated with socioeconomic, geographic, maternal and child characteristics

The inequities in basic vaccination coverage in Ethiopia associated with socioeconomic, geographic, maternal and child characteristics are illustrated in Table 4 and Figure 4. Children in the richest wealth quintile households had 3 times more likely to have basic vaccination coverage compared to children in the poorest wealth quintile households (adjusted OR 3.41; 95% CI: 1.50-7.73). Children in Addis Ababa and Dire Dawa were 11 times more likely to have basic vaccination coverage compared to children living in the Afar region (adjusted OR 11.60 and 11.62; 95% CI: 3.75-35.92 and 4.49-30.10 respectively). Children whose mothers had received primary education were 54% more likely to have basic vaccination coverage compared to children whose mothers had no education (adjusted OR 1.54; 95% CI: 1.08-2.19). Children living in a household headed by a woman were 48% less likely to have full basic vaccination coverage compared to households headed by a man (adjusted OR 0.52; 95% CI: 0.35-0.79).

**Table 4:** Inequities in basic vaccination coverage in Ethiopia associated with socioeconomic, geographic, maternal and child characteristics. Inequities in basic vaccination (1-dose BCG, 3-doses DPT-HepB-Hib, 3-doses polio, 1-dose measles vaccine) coverage among children aged 12-23 months associated with socioeconomic (household wealth), geographic (area of residence, region), maternal (maternal age at birth, maternal education, sex of household head) and child (sex of child, birth order) characteristics, based on multiple logistic regression estimates of adjusted odds ratios (mean and 95% confidence intervals).

|  | Adjusted Odds Ratios | p-value |
| --- | --- | --- |
| **Household wealth** (quintiles) |  |  |
| poorest (referent) |  |  |
| poorer | **1.91** (1.15, 3.18) | 0.01 |
| middle | **1.86** (1.07, 3.26) | 0.03 |
| richer | **2.45** (1.42, 4.20) | 0.001 |
| richest | **3.41** (1.50, 7.73) | 0.003 |
| **Area of residence** |  |  |
| rural (referent) |  |  |
| urban | 1.01 (0.39, 2.61) | 0.99 |
| **Region** |  |  |
| Afar (referent) |  |  |
| Tigray | **8.52** (3.43, 21.17) | <0.001 |
| Amhara | **3.30** (1.35, 8.07) | 0.009 |
| Oromia | 1.30 (0.54, 3.10) | 0.56 |
| Somali | 1.91 (0.75, 4.84) | 0.17 |
| Benishangul | **7.40** (3.02, 18.14) | <0.001 |
| Southern Nations,<br>Nationalities & People | **3.18** (1.29, 7.83) | 0.01 |
| Gambela | 2.32 (0.93, 5.77) | 0.07 |
| Harari | 1.89 (0.78, 4.54) | 0.16 |
| Addis Ababa | **11.60** (3.75, 35.92) | <0.001 |
| Dire Dawa | **11.62** (4.49, 30.10) | <0.001 |
| **Maternal age at birth** (years) |  |  |
| 15-19 (referent) |  |  |
| 20-34 | 1.64 (0.97, 2.78) | 0.06 |
| 35-49 | 1.20 (0.62, 2.34) | 0.60 |
| **Maternal education** |  |  |
| none (referent) |  |  |
| primary | **1.54** (1.08, 2.19) | 0.02 |
| secondary/higher | 2.12 (0.92, 4.88) | 0.08 |
| **Sex of household head** |  |  |
| male (referent) |  |  |
| female | **0.52** (0.35, 0.79) | 0.002 |
| **Sex of child** |  |  |
| male (referent) |  |  |
| female | 1.11 (0.82, 1.51) | 0.50 |
| **Birth order** |  |  |
| 6<sup>th</sup> born or higher (referent) |  |  |
| First born | 1.29 (0.71, 2.34) | 0.41 |
| 2<sup>nd</sup>-3<sup>rd</sup> born | 1.18 (0.73, 1.92) | 0.49 |
| 4<sup>th</sup>-5<sup>th</sup> born | 1.17 (0.72, 1.90) | 0.51 |

### Ethiopia Mini DHS 2019 update

In the report on the Ethiopia Mini Demographic and Health Survey 2019 [12], data from vaccination card, mother’s report and health facility records were collected for 1026 children aged 12-23 months for status on age-appropriate vaccinations (1-dose BCG, 3-doses DPT-HepB-Hib, 3-doses polio, 1-dose measles vaccines, 3-doses PCV and 2-doses rotavirus vaccines), and 1028 children aged 24-35 months for status on the second dose of measles vaccination (MCV-2). The mean DTP3 vaccination coverage was 60.9%, and vaccination coverage for other single vaccines ranged from 58.6% for MCV-1 to 73% for BCG vaccine, and 9.1% for MCV-2 which was launched in early 2019. The vaccination coverage for basic vaccinations (1-dose BCG, 3-doses DPT-HepB-Hib, 3-doses polio, 1-dose measles vaccines) was 43.1%, while coverage for 3-doses PCV and 2-doses rotavirus vaccines were 59.9% and 66.7% respectively.

The basic vaccination coverage was relatively higher at 57.3% in urban areas in comparison to rural areas at 36.9%, and 83.3% and 18.2% in Addis Ababa and Somali regions respectively. The basic vaccination coverage was also higher among children from richer households and higher maternal education with 64.7% and 24.7% in the highest and lowest wealth quintiles respectively, and 65.1% and 33.8% among children of mothers with more than secondary education and no education respectively.

## Discussion

### Main findings

Household wealth status, administrative regions, maternal education and sex of household were identified to be strongly associated with basic vaccination coverage (1-dose BCG, 3-doses DPT-HepB-Hib, 3-doses polio, 1-dose measles vaccine) after adjusting for background characteristics. Higher levels of basic vaccination coverage were associated with children from richer households, urban regions of Addis Ababa and Dire Dawa, primary maternal education and male headed households in comparison to children from poorer households, rural regions of Afar and Somali, no maternal education and female headed households respectively.

The Ethiopian National Expanded Programme on Immunisation and the Global Vaccine Action Plan had set coverage goals of 90% at national level and 80% at district level by 2020 [4,13]. But, basic vaccination coverage (1-dose BCG, 3-doses DPT-HepB-Hib, 3-doses polio, 1-dose measles vaccine) in Ethiopia was well below the coverage goals at 39.7% and 43.1% in 2016 and 2019 respectively. The coverage of single vaccines range between 53.2% and 69.2% in 2016 and between 58.6% and 73% in 2019.

### Health card and vaccination records

It was found that 57.5% (1152/2004) of children age 12-23 months in 2016 had a health card or a vaccination record in a health facility at the time of the interview, and 64.5% (743/1152) of these children had full basic immunisation coverage. Of the remaining 42.5% (852/2004) without a vaccination record or health card, only 6.2% (53/852) had basic immunisation coverage. Thereby, lack of vaccination records and health records signal a bottleneck for immunisation access, and measures to resolve this bottleneck are likely to include these missing children within the immunisation system and improve vaccination coverage.

### Geographical factors

Nearly half of the contributing factors for under-immunisation in low- and middle income countries are immunisation systems related, with geographical factors affecting access and distance to health facilities being most common [23]. There are large differences in odds of basic vaccination coverage according to regions in this study which adds evidence towards geographical barriers to accessing immunisation services. Low population density and weak health infrastructure have been identified as challenges for routine immunisation services in Afar, Somali and Gambella [24]. Afar and Somali regions in particular are dominated by pastoralist nomadic communities. Considering weak health infrastructure, distance to health facilities and mobility patterns of the population, routine immunisation services are difficult to deliver. Consequently, the current comprehensive National Immunisation Plan for 2016-2020 identifies pastoralist communities as populations at risk of missing immunisation services. An example of an outreach approach are pre-arranged locations and dates for government health workers to meet with nomadic communities where vaccinations and basic health services are provided as well as scaling up the quantity of services [13,24]. Thereby, improved understanding of spatiotemporal nomadic patterns might facilitate the service delivery of these outreach programmes to improve coverage and equitable uptake of vaccines among pastoralist communities. Also, taking into account the wide reach of immunisation programmes, improving equity in immunisation coverage through improved service delivery and outreach may simultaneously improve the delivery of other health interventions and preventive measures focused on child health [25].

### Maternal education

Maternal education was associated with improved vaccination coverage in this study and similar findings have been described for Ethiopia, in multi-country assessments of inequities in vaccination coverage, and in systematic reviews of studies from 13 and 51 countries [9,23,26–30]. A higher maternal educational level raises awareness about the importance of vaccination and it has been reported that educated women choose health care services that generate better health [27,31].

### Household wealth status

While household wealth was associated with improved vaccination coverage, vaccinations are provided free of charge at the public health facilities in Ethiopia. Thereby, there are no financial barriers in the form of out of pocket-payments for immunisation services and political efforts have been made to reach rural and poor population groups through health extension workers based in communities [13,24,32]. Multi-country studies have found associations of children from households of lower socioeconomic status with lower vaccination coverage [23,26,29,33]. In Ethiopia, there are likely to be additional financial barriers beyond out of pocket-payments such as transportation costs and productivity loss from taking time off to access vaccination services and other non-financial barriers to vaccine acceptance among lower socioeconomic households, which warrants additional studies to explore and identify the underlying causal mechanisms.

### Gender of household head

Children living in female headed households headed were less likely to have basic vaccination coverage in Ethiopia. A systematic review on the influence of women’s empowerment on basic immunisation coverage concluded that women’s agency measured as decision-making capacity is positively associated with their children’s vaccination coverage [34]. The contrary findings with lower vaccination coverage among children living in female headed households in Ethiopia warrant further exploration into associations between female agency and health-related decision-making. Sociocultural aspects relating to marital status and higher agency of married women living with their partner could be a potential explanation for the contrary findings.

### Immunisation services

The differences in coverage between basic vaccination and single vaccinations can be partly explained by factors relating to immunisation services and missed opportunities to vaccinate. Within the three-tiered primary health care system, immunisation services are mainly delivered by health officers (nurses) who are based in health centres, health extension workers based at health posts which are satellite posts of health centres, and a volunteer workforce of health development army. Low monitoring and supervision capacities prevent a focus on immunisation service delivery and result in insufficient feedback given to health staff. Despite a government policy requiring health facilities to offer immunisation services on a daily basis, a 2015 survey found that only 51% of health centres and 11% of health posts offered daily immunisation services in Ethiopia due to shortage of qualified health workers and challenges relating to vaccine supply and cold chain management [35]. 31% of health facilities had refrigerators, but only 11% had adequate refrigerator temperature. The cold chain rehabilitation and expansion plan aims to address these challenges by improving both logistic and storage aspects of vaccine distribution [24].

### Missed opportunities for vaccination

Despite contact with health services, there are additional missed opportunities for vaccination. More than 50% of mothers and children in Ethiopia received one or more health interventions of antenatal care, postnatal care, vitamin A supplement, skilled birth attendants and insecticide-treated bed-nets but have failed to have their children fully vaccinated [36]. Missed opportunities to administer all vaccines scheduled at the same visit and thus resulting in a low basic vaccination coverage, can indicate difficulties in vaccine supply, may relate to health workers’ reluctance to administer multiple vaccines simultaneously or to knowledge gaps in identifying which vaccines are due [37]. Supplementary immunisation activities for polio, measles, tetanus and meningococcal A vaccinations during 2011-2014 [13], which are mass-immunisation campaigns targeted at children otherwise missed by routine services, could lead to a one-sided focus on specific vaccines while missing out on other routine vaccinations [38].

### Related studies on vaccination coverage and equity impact in Ethiopia

In the survey analysis study by Tamirat et al on full immunisation coverage and associated factors among children aged 12-23 months in Ethiopia using DHS 2016, female headed households and rural dwellings were negatively associated with basic vaccination while higher maternal education, employment, middle and rich economic status, ante-natal care follow up, and delivery at health facility were positively associated with basic vaccination [39]. In the community-based cross-sectional study by Marefiaw et al on age-appropriate vaccination and associated factors for pentavalent and measles vaccination in Menz Lalo district of northeast Ethiopia, age-inappropriate pentavalent 1-3 vaccinations was associated with being male sex, lack of telephone, lack of usual caretaker, unplanned pregnancy, missing pregnant women’s conference, decreasing birth order and insufficient knowledge [40]. In the spatial and multilevel analysis study by Geremew et al on the geographical variation and associated factors of childhood measles vaccination in Ethiopia, seven significant space clusters with low MCV1 coverage were detected, with the primary, secondary and tertiary clusters located in the Afar, Somali and Gambella regions respectively [41]. Child age, first and third doses of pentavalent vaccination, secondary and above maternal education, and media exposure increased the odds of MCV1 vaccination while children with older maternal age had lower odds of MCV1 vaccination. Finally, there are differences in vaccination coverage estimates between DHS, WUENIC and official estimates of national authorities that should be taken into account in guiding policy and practice to improve vaccination coverage and equity in Ethiopia (see Appendix A3).

### Gavi support for Ethiopia

A goal of Gavi’s 2021-2025 strategy is “Gavi will bring a much stronger focus on reaching those most marginalised, by strengthening primary health care systems, building and sustaining community demand, and using innovation to ensure that immunisation services reach these children”, which is in line with the ‘leave no one behind’ and ‘reaching the furthest behind first’ principles of the SDGs [6,42]. In a situation where Ethiopia is facing a phase-out of donor support for several programmes which are aimed at improving vaccination coverage in underserved administrative regions and zones, such as the Routine Immunisation Improvement Plan [13,24], such a focus and subsequent support by Gavi could be crucial in improving vaccination coverage among the children who are currently not reached by immunisation services.

## Conclusion

In the comprehensive multi-year plan 2016-2020 of the Ethiopia National Expanded Programme on Immunisation, the Ministry of Health identifies inequities in access to immunisation services, as well as regional disparities, as major weaknesses in immunisation service delivery. The Ministry of Health has proposed to tackle these inequities by implementing all components of the WHO/UNICEF “Reaching every community” approach during 2016-2020 [13]. This study contributes to this approach by analysing childhood vaccination coverage in Ethiopia by socioeconomic, geographic, maternal and child characteristics to identify population subgroups with low vaccination coverage -- children from poorer households, rural regions of Afar and Somali, no maternal education and female headed households. These findings are aimed to facilitate the design and development of targeted approaches that are adapted to the local context of socioeconomic, geographic, maternal and child characteristics to improve vaccination coverage and equitable uptake of vaccines in Ethiopia.

## Data Availability

The 2016 Ethiopia DHS data file (ETKR71DT.ZIP) used in this study is accessible on the DHS website (https://dhsprogram.com/data/dataset/Ethiopia_Standard-DHS_2016.cfm) and available to registered users

## Acknowledgements

We had presented the study results to the Ethiopia Interest Group at the London School of Hygiene and Tropical Medicine and we thank them for their valuable feedback. We thank Schadrac Agbla, Ana Maria Buller, Lars Åke Persson, Billy Quilty and Anne Schlotheuber for helpful discussions. KMA is supported by NIH/NIGMS R01GM109718 and the Vaccine Impact Modelling Consortium (OPP1157270).

## Appendix

### Appendix A1. Model development and variable selection for multiple logistic regression

Simple logistic regression was conducted to assess associations of basic vaccination coverage with socioeconomic (household wealth, religion, ethnicity), geographic (area of residence, region), maternal (maternal age at birth, maternal education, maternal marital status, sex of household head) and child (sex of child, birth order) characteristics respectively.

Causal modelling through multiple logistic regression was then conducted to assess adjusted associations between basic vaccination coverage and socioeconomic (household wealth), geographic (area of residence, region), maternal (maternal age at birth, maternal education, sex of household head) and child (sex of child, birth order) characteristics. Maternal marital status was excluded in the multiple logistic regression analysis due to collinearity with sex of household head -- in households where married women are living with a partner, household head was likely to be male. Religion and ethnicity were excluded from the multiple logistic regression analysis due to collinearity with region -- administrative regions in Ethiopia are inhabited by major ethnic groups specific to that region and the predominant religions are specific to the ethnic groups.

Joint hypothesis testing using a Wald test was performed for categorical exposure variables with two or more subgroups to test the null hypothesis of no overall association between the variable and basic immunisation coverage. In a final step, effect modification between selected exposure variables and basic vaccination coverage was investigated by including interaction terms in the regression model. Interaction between:

- household economic status (i.e. wealth quintile) and area of residence
- household economic status and maternal age
- household economic status and maternal education
- maternal age and maternal education

was tested using the *contrast* command in Stata. Due to small sample size in some subgroups, the exposure groups of maternal education were combined to create a binary variable and thus increase the power of testing for interaction

### Appendix A2. Reproducible analysis

The 2016 Ethiopia DHS data file (ETKR71DT.ZIP) used in this study is accessible on the DHS website (https://dhsprogram.com/data/dataset/Ethiopia_Standard-DHS_2016.cfm) and available to registered users. The Stata codes used for the study analysis are available upon request from the corresponding author.

### Appendix A3. Vaccination coverage estimates for Ethiopia

There are differences in vaccination coverage estimates for 2016 between Ethiopia DHS, WUENIC (WHO/UNICEF Estimates of National Immunization Coverage) and official estimates of national authorities, as shown in Table A3.1. DHS is based on population-based household surveys among children aged 12-23 months based on the information on vaccination history from documented evidence or caregiver recall. WUENIC uses both administrative as well as survey data, which are assessed for potential biases and the final estimates are only derived after consultation with local partners in the respective countries [16,43]. Official estimates reported by national authorities reflect their assessment based on administrative coverage, survey-based estimates or other data sources or adjustments. The differences in vaccination coverage estimates between different sources should be taken into account in guiding policy and practice to improve vaccination coverage and equity in Ethiopia.

**Table A3.1.** DHS, WUENIC and official estimates of vaccination coverage in Ethiopia.

| **Vaccine and dose** | **2016 DHS**<br>**(children aged**<br>**12-23 months)** | **WUENIC 2014**<br>**estimates**<br>**(unadjusted)** | **WUENIC 2014**<br>**estimates**<br>**(adjusted)** | **Official country**<br>**estimates (2014)** |
| --- | --- | --- | --- | --- |
| BCG | 69 | 75 | 69 | 78 |
| DTP 1 | 73 | 86 | 73 | 89 |
| DTP 3 | 53 | 77 | 61 | 84 |
| Polio 3 | 56 | 75 | 68 | 84 |
| Measles 1 | 54 | 70 | 54 | 81 |
| PCV 3 | 49 | 76 | 55 | 82 |
| Rotavirus 2 | 56 | 63 | 56 | 70 |

## Notes

### Competing Interest Statement

The authors have declared no competing interest.

### Clinical Trial

not applicable

### Funding Statement

Anne Geweniger received no external funding.
Kaja M. Abbas is supported by NIH/NIGMS R01GM109718 and the Vaccine Impact Modelling Consortium (OPP1157270).

